# COMMUTE: communication-efficient transfer learning for multi-site risk prediction

**DOI:** 10.1101/2022.03.23.22272834

**Authors:** Tian Gu, Phil H Lee, Rui Duan

## Abstract

**Objectives:** We propose a communication-efficient transfer learning approach (COMMUTE) that efficiently and effectively incorporates multi-site healthcare data for training risk prediction models in a target population of interest, accounting for challenges including population heterogeneity and data sharing constraints across sites.

**Methods:** We first train population-specific source models locally within each institution. Using data from a given target population, COMMUTE learns a calibration term for each source model, which adjusts for potential data heterogeneity through flexible distance-based regularizations. In a centralized setting where multi-site data can be directly pooled, all data are combined to train the target model after calibration. When individual-level data are not shareable in some sites, COMMUTE requests only the locally trained models from these sites, with which, COMMUTE generates heterogeneity-adjusted synthetic data for training the target model. We evaluate COMMUTE via extensive simulation studies and an application to multi-site data from the electronic Medical Records and Genomics (eMERGE) Network to predict extreme obesity.

**Results:** Simulation studies show that COMMUTE outperforms methods without adjusting for population heterogeneity and methods trained in a single population over a broad spectrum of settings. Using eMERGE data, COMMUTE achieves an area under the receiver operating characteristic curve (AUC) around 0.80, which outperforms other benchmark methods with AUC ranging from 0.51 to 0.70.

**Conclusion:** COMMUTE improves the risk prediction in the target population and safeguards against negative transfer when some source populations are highly different from the target. In a federated setting, it is highly communication efficient as it only requires each site to share model parameter estimates once, and no iterative communication or higher-order terms are needed.

## Introduction

Risk prediction models play an important role in precision medicine and clinical care, showing great potential to assist clinical decision-making and enhance the quality of care delivered to patients^17^. In recent years, growing efforts have been devoted to constructing institutional or national biobanks, where individuals’ electronic health records (EHR) are linked with genomics, imaging, and behavioral observations. For example, the UK Biobank is a prospective cohort of over 500,000 samples with genetic data and health information^49^. In the United States, institutional biobanks such as Bio*Me* from the Icahn School of Medicine at Mount Sinai^1,4^, BioVU at Vanderbilt University^41,46^, and the Mass General Brigham (MGB) Biobank^26^ enroll increasing numbers of participants every year to measure their genetic variants, biomarkers, metabolic data and a large number of disease phenotypes. These datasets are valuable resources to assess individuals’ risks of developing common and many complex diseases.

Despite the availability of multiple biobanks, one common challenge we face is the diminished performance when applying models trained in one dataset to a different dataset, which is known as the lack of model transferability and portability^18^. The heterogeneity in the underlying distribution of data across different populations may explain such low portability. For example, compared to many healthcare center-based biobanks, the UK Biobank has older participants with lower disease prevalence across many disease categories^3^. Another example is the lack of transferability of genetic risk prediction models across ethnic groups^13,34^, resulting from the substantial heterogeneity in the genetic architectures, linkage disequilibrium, and allele frequency^13,34^.

Practically, it is often of interest to train a risk prediction model for a particular population (referred to as the target population hereafter), for example, a demographic sub-population or one of the healthcare centers in a research network. Due to the lack of portability, a population-specific training strategy is commonly used, which might require a substantial amount of data from the target population to obtain relatively good model performance. However, training a model using only the target data may lead to unsatisfactory performance in cases where limited data are observed from the target population. For example, the target population may be an underrepresented demographic sub-population or a healthcare center with limited labeled data. Another strategy is to pool all data together and train a unified model across all populations, in which case the sample size is increased, but it fails to adjust for population-level heterogeneity. In other words, a model trained in such a way is not particularly representative of the target population of interest but of the pooled overall population. Such a unified training strategy might be problematic in cases where the target population is underrepresented and potentially has different distributional characteristics from the overall population. Methodologically, it is still a challenge to effectively and efficiently incorporate heterogeneous data towards risk prediction in a target population while addressing other practical issues such as data sharing constraints.

Regarding data sharing, some research networks adopt a centralized model where multi-site individual-level data are de-identified and stored in a central data warehouse^16,19,22,37^. Although a centralized model is ideal for analyzing multi-site data, it is less feasible in many settings due to data privacy, storage, and management barriers^25^. Many networks, therefore, employ a federated model where data remain locally in each site while only summary-level statistics are shared across sites for collaborative data analysis^5,24,48,64^. The federated model helps construct a highly collaborative community that attracts more participating sites and enables rapid development and deployment of research projects across institutions. In addition, we also see the success of some flexible hybrid models, especially when collaborations are built across networks^56^. Consequently, flexible methods that can be efficiently implemented in different research networks are needed.

To overcome data sharing barriers, researchers developed distributed algorithms based on summary statistics for jointly training models across multiple datasets without sharing individual-level data. For example, a series of algorithms were developed for regression models by sharing gradients of objective functions^11,12,33,42,59^. Other approaches involve sharing parameter estimates fitted locally at each site^7,28,63^. However, most of the aforementioned work aims to train a unified model across multiple datasets, ignoring heterogeneity across datasets. Federated learning methods were developed for collecting and updating models between each client’s devices and a central server, which allows client-specific model components to account for differences in each client’s data^40^. Nevertheless, these approaches often require iterative model updating across devices and the central server^32,33,47^, which may be impracticable in research networks or cross-network collaborations without required infrastructures for timely and efficient model updating. In many research networks, iterative communication across sites requires extensive effort and time. Thus, there is a great need for communication-efficient algorithms that only require each site to share summary statistics once^11,12,42^.

In this paper, we developed a communication-efficient transfer learning approach (COMMUTE) which provides a flexible and robust framework for multi-site risk prediction. COMMUTE can be applied in networks that employ centralized, federated, or hybrid data sharing models with at most one round of communication across sites. Instead of a site-specific or a unified training strategy, COMMUTE is built on a transfer learning algorithm^6,31,38,58^, which adjusts for between-population heterogeneity through distance-based adaptive regularization. Our data-driven method avoids negative transfer from sites that are substantially different from the target population, and therefore it is robust to the level of heterogeneity across populations^54,57^.

## Methods

### Notation and problem setup

We consider the setting where there are multi-site data collected from *K* + 1 populations, including one target population of interest and *K* source populations that are potentially helpful to the training of a prediction model in the target population (indexed by *k* = 1, …, *K*). We consider a general situation, where among the *K* source datasets, *M* can be directly combined with the target dataset, and the remaining *K* − *M* datasets have data-sharing constraints that the individual-level data can only be analyzed locally and only the summary-level information is shareable. Note that *M* = 0 indicates a completely federated setting; 0 *< M < K* indicates a hybrid setting where partial data can be pooled; and *M* = *K* indicates a centralized setting where all data are stored together. The target data has sample size *n* and the *k*-th dataset has sample size *n*_*k*_, where the total sample size is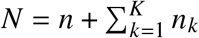.

In the target population, we have the outcome variable *Y* ∈ ℝ^*n*^ and a set of *p* predictors *X* ∈ ℝ^*n*×*p*^, which follows some distribution, denoted by *f* (*y*, **x**). We consider a generic class of machinelearning methods where fitting the model involves a loss function indexed by a set of parameters, denoted by *L* (**b**; *Y, X*) with **b** ∈ ℝ^*m*^. For example, in the generalized linear models (GLM), *L* (**b**; *Y, X*) is the negative log-likelihood function, and **b** corresponds to the regression coefficients and over-dispersion parameters^35^; when using support vector machine (SVM), *L* (**b**; *Y, X*) could be the Hinge loss function and **b** represents the parameters of the kernel function^9^. We define the target parameter *β* to be

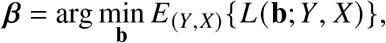

where the expectation is taken with respect to the distribution of the target data. With the target data alone, we can obtain a *target-only* estimator by optimizing the target loss function, i.e.,

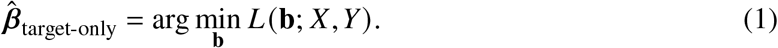

which might be improved when borrowing information from available source populations.

For the *k*-th source population, we observe the outcome variable 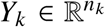 and the same set of *p*-dimensional predictors 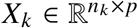 as in the target population. Similarly, define

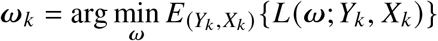

to be the source parameters, where the expectation is taken with respect to the distribution of data at the *k*-th population, i.e., *f*_*k*_ (*y*, **x**). Due to population heterogeneity, the parameters 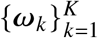 and *β* can be different. On the other hand, they may share certain similarities in their structures or magnitudes such that information about *ω*_*k*_ could guide the learning of *β*. Our goal is to leverage such similarities in a data-driven way to improve the estimation of *β*, overcoming additional challenges of data sharing.

To achieve this goal, we first need to characterize the similarity between the source and the target parameters. In previous work, such as Li et al. ^30^, Tian and Feng ^52^, Xu and Bastani ^60^, it is assumed that a source population *k* is helpful to the target when the difference *β* − *ω*_*k*_, is a sparse vector. That is saying, most of the model parameters take the same value across populations, while only a small subset of the parameters may take different values. A similar idea has been seen in many network-based deep transfer learning methods where the target data are used to selectively retrain (fine-tuning) some of the layers of pre-trained neural network models from a source population^50^. In comparison to the existing work, our method allows more flexibility when characterizing the similarity between *β* and *ω*_*k*_. Intuitively, if there exists some distance measure *d* (*β, ω*_*k*_) which is small, *ω*_*k*_ can be leveraged to provide additional information in the estimation of *β* through imposing a similarity constraint *d* (*β, ω*_*k*_) ≤ *h*. The smaller *h* is, the source data would be more helpful to the training of the target model as the parameter space is reduced by the similarity constraint. In our work, we propose a flexible distance measure defined as *d* (*β, ω*_*k*_) = ∥ *β* − *ω*_*k*_ ∥_*q*_, where *q* can take any positive numbers. Choosing *q* = 0 leads to the similarity measure considered in Li et al. ^30^,^31^, Tian and Feng ^52^, which quantifies how many parameters have identical values across two populations. When *q* = 2, it represents the Euclidean distance characterizing the differences over all parameters. When ∥ *β* − *w*_*k*_ ∥_2_ is small, it allows *β* − *ω*_*k*_ to be a non-sparse vector but the magnitude of the differences cannot be too big. In practice, one can flexibly choose *q* to better capture the differences in the model parameters across populations, which can be prespecified if prior knowledge on the structure of *β* − *ω*_*k*_ is known, or it can be determined in a data-driven way through cross validation. In addition, weighted distance measures can be designed if prior knowledge is available regarding which parameters are more likely to be similar across two populations.

### Communication-efficient transfer learning (COMMUTE)

Our proposed approach, COMMUTE, involves three main steps (see Figure 1). In the following, we keep a generic form of the risk prediction model, and use a penalized logistic regression model as an example to introduce the method.

**Figure 1:**
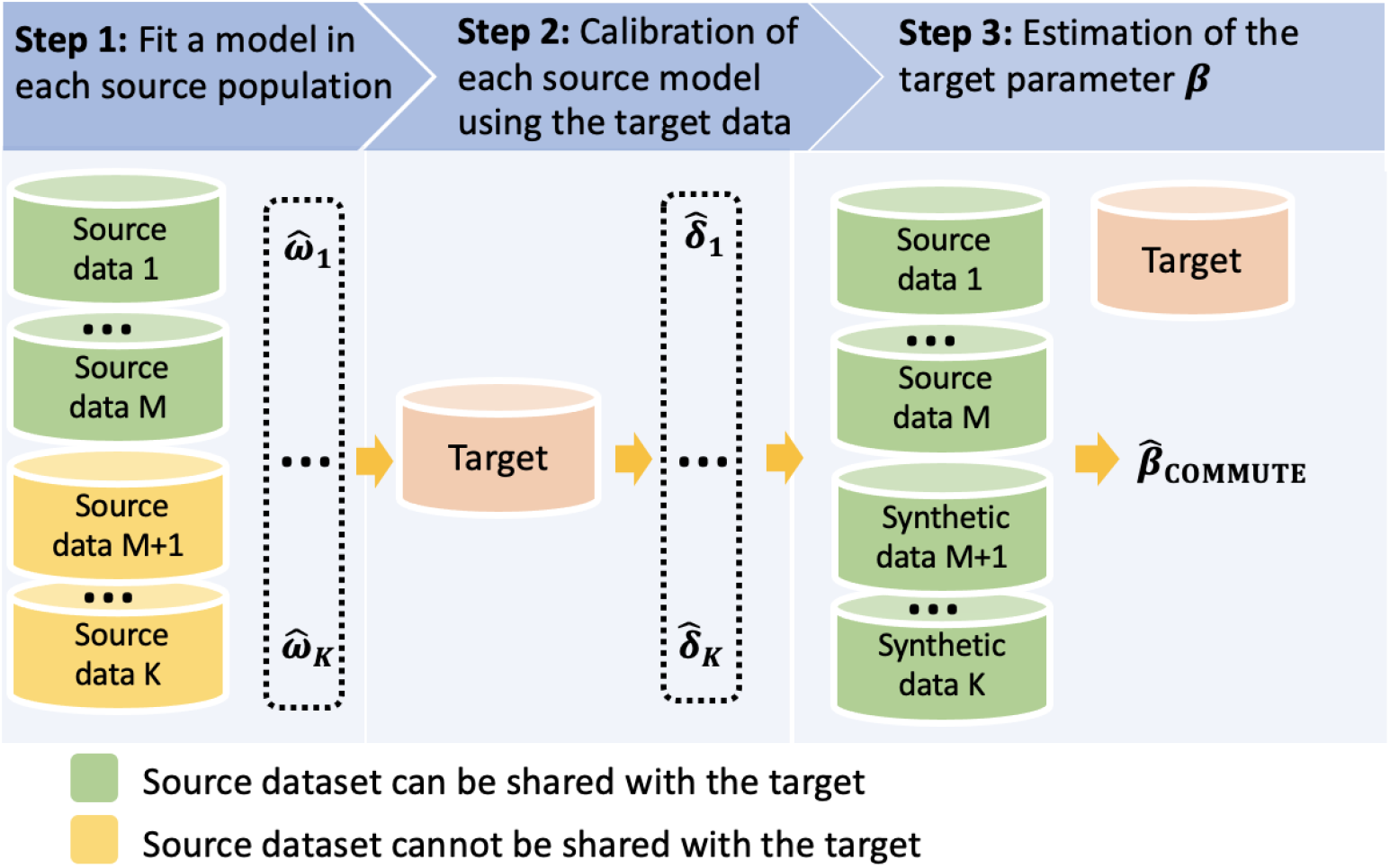
Workflow of COMMUTE.

#### Step 1: Fit a model using data from each source population

The source-specific parameter estimate from the *k*-th site, denoted by 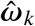, can be obtained by minimizing the loss function *L* (*ω*; *X*_*k*_, *Y*_*k*_) determined by the risk prediction method. In the existence of additional assumptions on the model parameters, e.g., sparsity or other structured patterns, additional penalty terms or constraints can be added to the optimization. A generic form is summarized as

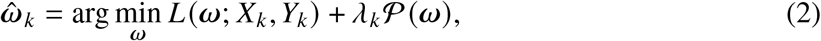

where 𝒫(·) is some penalty term (added when necessary) and, *λ*_*k*_ is a tuning parameter that can be selected through cross validation. Using a *L*_1_ penalized logistic regression as an example, one can take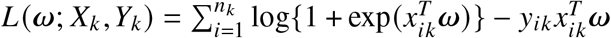, where *x*_*ik*_ are the predictors including the intercept term for the -th individual in the *k*-th site, *y*_*k*_ is the outcome, and 𝒫(*ω*) = ∥ *ω* ∥_1_. In the case where individual-level data from the *k*-th source population cannot be shared with the target, only the source estimator 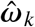 needs to be shared with the target site, a vector containing *m* numbers.

#### Step 2: Calibration of each source model using data from the target population

In this step, we use the target data to estimate the parameter difference, denoted as *δ*_*k*_ = *β* − *ω*_*k*_. Specifically, we can learn *δ*_*k*_ through

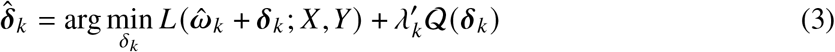

where 𝒬(*δ*_*k*_) is a penalty term depending on the similarity measure *d* (*β, ω*_*k*_) = ∥ *β* −*w*_*k*_ ∥_*q*_ = 1*δ*_*k*_ 1_*q*_ and, 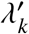 is a tuning parameter. When *d* (*β, ω*_*k*_) is small, the source estimator 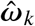 can help reduce the search space of the target parameters through the penalization on *δ*_*k*_. For example, if we believe that *d* (*β, ω*_*k*_) = ∥*δ*_*k*_ ∥_0_ is small, we may choose 𝒬(*δ*_*k*_) = ∥*δ*_*k*_ ∥_0_; or we may choose Q(*δ*_*k*_) = ∥*δ*_*k*_ ∥_1_, a convex relaxation of the *L*_0_ penalty that also leads to sparse solutions^53^. With such penalization, the source estimator 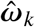 is used to guide the learning of the target parameter *β*. This step is performed within the target site, so no extra communication across sites is needed.

#### Step 3: Estimation of the target parameter ***β***

To estimate *β*, in the centralized setting, i.e., *M* = *K*, we propose a transfer learning estimator, defined as

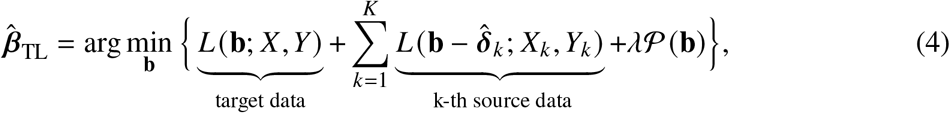

where, *λ* is a tuning parameter. In this joint estimation, the calibration term 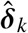from Step 2 is used to adjust for the difference between the *k*-th source and the target. In comparison to training a unified model using all the source and target data, i.e.,

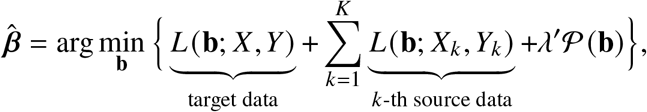

where, *λ* ^′^ is a tuning parameter, the unified model ignores the fact that *β* is different from 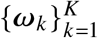. Imagine that the *k*-th source population has a much larger sample size than other populations, 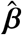 will likely be close to *ω*_*k*_, instead of *β*.

In the federated or hybrid setting, i.e., 0 ≤ *M < K*, we cannot obtain 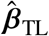, since individual-level data from *K* − *M* populations cannot be pooled with the rest of the *M* datasets. To tackle this challenge, we propose to generate *K* − *M* sets of heterogeneity-adjusted synthetic data in the target site. These synthetic datasets will then be combined with the target data and the *M* source datasets to jointly estimate *β*.

Specifically, without loss of generality, we assume that the first *M* source datasets, denoted by *k* = 1, …, *M*, can be shared with the target. For each *k* ∈ {*M* + 1, …, *K* }, we have obtained their source estimator 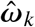 and the calibration term 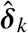 from Steps 1-2. Using 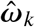 and 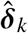, we can compute 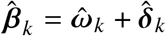, an estimator for the target parameter *β* that only incorporates information from the *k*-th source. With some positive integer *r*, we then generate a synthetic dataset with a sample size *n*_*k*_ × *r*, where the *p* predictors are obtained by random sampling from the target *X* (denoted by 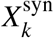), and the outcome (denoted by 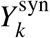) is generated by applying the prediction model with model parameter 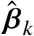.

Finally, the COMMUTE estimator can be obtained by jointly analyzing a combined data of the target, the source and the synthetic data:

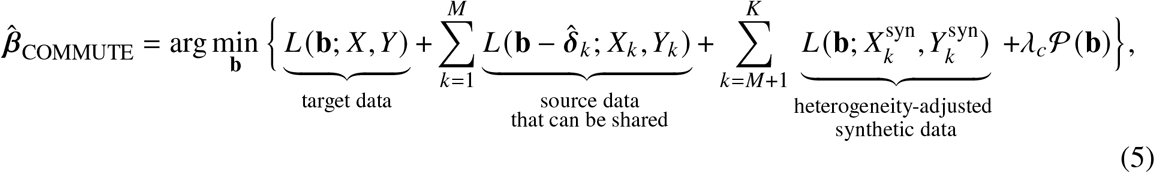

where, *λ*_*c*_ is a tuning parameter. When 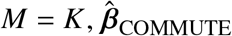 is the same as 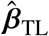. A detailed algorithm summarizing Steps 1 to 3 is shown in Algorithm 1.

##### Algorithm 1: the COMMUTE algorithm

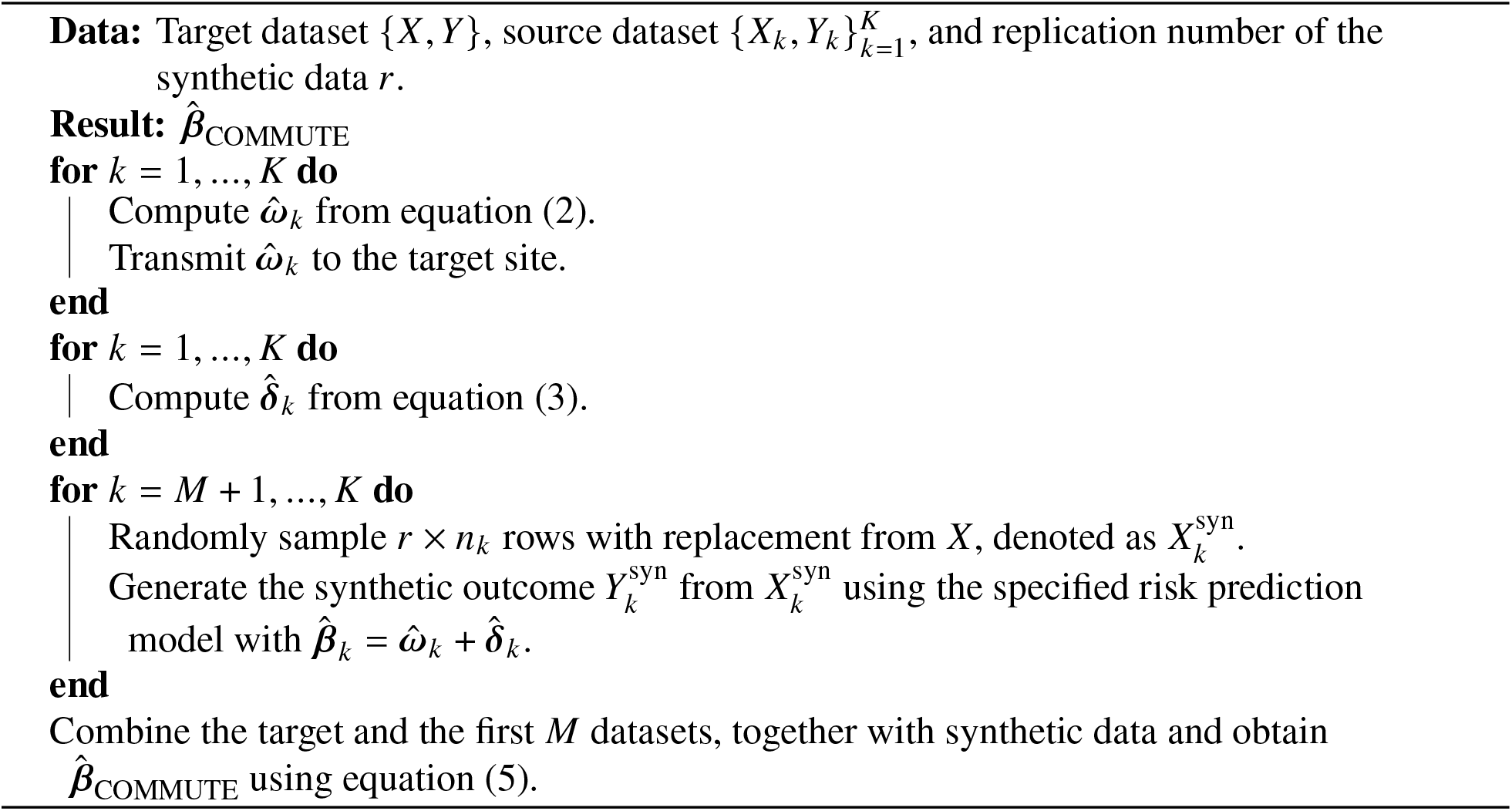

Similar ideas of creating synthetic data from summary statistics of external studies have been considered in association studies20,21, survey methodologies^43,44,45^ and causal inference^51^. Compared to those methods, our approach allows heterogeneous underlying distributions and corrects such cross-population heterogeneity before creating synthetic data. Compared to existing federated learning methods that require iterative model updating, COMMUTE is more communication-efficient as it only requires one round of communication for the source sites to share the model estimates with the target site. Compare with methods that protect data privacy by generating and sharing synthetic data which mimic the distribution of the original data^8,10,39^, we directly use re-sampling since our synthetic data is generated within the target site (not to be shared across sites) and privacy is therefore not a concern.

The optimal choice of *r* depends on the similarity between the target and the source populations. As our numerical investigation suggests (See Figure S1 in the Supplementary Material), when the discrepancy between the source and target parameters is low or moderate, increasing *r* improves the performance of COMMUTE. When the difference is large, increasing *r* decreases the performance. In both cases, increasing *r* reduces the gap in the performance between the federated version of COMMUTEand the centralized version of COMMUTE 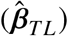, and the performance of COMMUTE becomes stable as *r* continues to increase. In practice, if computationally allowed, we suggest choosing a large *r*, assuming the source populations are helpful. Since this assumption might not be true and some source populations might be highly different from the target, we introduce an additional step to prevent negative transfer.

### A safeguard to prevent negative transfer

When some of the source populations are highly different from the target population, including the source may lead to performance worse than not including it. To prevent negative transfer, we propose to first rank the *K* sites by the performance of source estimators 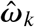 on the target training data. We can then only include sites with performance higher than a certain threshold which can be either pre-specified or determined by cross-validation. Another way is to sequentially incorporate populations based on their rankings. For example, we set the first estimator as the target-only estimator 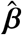, which does not include any source population. We can then only include the site whose model has the best performance on the target training data, and apply Algorithm 1 to obtain 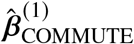. Sequentially including more source populations, we can further obtain 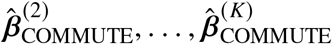. A small validation dataset can then be used to aggregate the models and obtain the final estimator that has the best performance among these candidate models^29,55^.

## Validation of COMMUTE

### Simulation studies

We evaluate the performance of COMMUTE using simulated data generated under different settings. We consider the case where we have a target site and three source sites. We generate predictors of dimension *p* = 2000 from multivariate Gaussian distributions with mean 0 and a population-specific covariance matrix Σ_*k*_ for the *k*-th population. We choose Σ_*k*_ to be a block-wise matrix with 20 blocks each of dimension 100 × 100 with an auto-regressive structure and the correlation was generated from a Uniform distribution between (0.2, 0.8).

With the simulated predictors, in the three source populations, we generate binary outcomes as Bernoulli random variables with probability Pr(*Y* |**x**) = expit(**x**^*T*^ *ω*_*k*_) for *k* = 1, 2, 3, respectively, and similarly in the target population we generate Bernoulli random variables with probability Pr(*Y* |**x**) = expit(**x**^*T*^ *β*). We set *β* to be sparse with 100 non-zero entries, which are generated from a Uniform distribution between (−0.5, 0.5). To generate *ω*_*k*_, we generate *δ*_*k*_ to be a sparse vector with *h*_*k*_ non-zero entries each with magnitude 0.5 and a randomly assigned sign from {−1, 1}. We then obtain *ω*_*k*_ = *β* − *δ*_*k*_. By increasing *h*_*k*_, we increase the number of parameters with different values from *β*, representing the increasing heterogeneity between the *k*-th source and the target population.

We evaluate the performance of the methods by varying the level of heterogeneity (*h*_*k*_) in each source population, as well as the sample size *n*_*k*_, with the target sample size *n* = 500. For the level of heterogeneity, we consider the following four settings:

(A) All three source sites have low-level heterogeneity, *h*_1_ = *h*_2_ = *h*_3_ = 20;

(B) All three source sites have moderate level heterogeneity, *h*_1_ = *h*_2_ = *h*_3_ = 60;

(C) All three source sites have high level heterogeneity, *h*_1_ = *h*_2_ = *h*_3_ = 120;

(D) Two source source have low-level heterogeneity, *h*_1_ = *h*_2_ = 20, and one has high level heterogeneity, *h*_3_ = 120.

We also consider two sample size settings:

1. *n*_1_ = 1000, *n*_2_ = 1500, *n*_3_ = 2000;

2. *n*_1_ = 2000, *n*_2_ = 3000, *n*_3_ = 4000.

We apply Algorithm 1 with the loss function of the logistic regression model. Given the sparse setting, we choose 𝒫(·) = 𝒬(·) = ∥ · ∥_1_. We choose *r* = 25. We compare the performance of the proposed method with another four methods: (i) target-only (Target): estimator obtained from equation (1); (ii) source estimators (Source): estimators obtained from equation (2); (iii) direct aggregation (Direct): we split the target data into training and validation with a ratio of 4:1, and we fit the target-only estimator using the training data, and use the validation data to learn a weighted average of the target-only and the source estimates where weights are proportional to the inverse of prediction errors evaluated in a validation data; and (iv) COMMUTE (Proposed): we consider four different cases: federated version (*M* = 0); hybrid version where only the first source datasets can be shared with the target (*M* = 1); hybrid version where the first two source datasets can be shared with the target (*M* = 2); centralized version where all data can be pooled together (*M* = 3). The same validation dataset as in the direct aggregation method is used for the additional step to prevent negative transfer (as introduced in the previous section). For each method, we calculate the average area under the operating characteristic curve (AUC) on an independent testing dataset of size 1000 over 200 iterations.

### Application to the prediction of extreme obesity using data from eMERGE

We evaluate the performance of COMMUTE using multi-site data from the eMERGE network obtained through the database of Genotypes and Phenotypes (dbGaP), where EHR-derived phenotypes for 55,029 subjects from ten participating sites were linked with DNA samples from biorepositories^19^. Among all subjects, 73% of the participants are White, 20% are African American (AA), 1% are Asian, and 6% are Unknown. We evaluate the performance of COMMUTE by constructing risk prediction models for extreme obesity using demographic features and genotypic data.

The extreme obesity phenotyping algorithm at eMERGE classifies each subject into either “Case”, “Control”, or “Neither Case nor Control”. We remove subjects with a “Neither Case nor Control” status and only include subjects with verified case-control status. Excluding three children’s hospitals, we initially obtain data from seven sites: Vanderbilt University, Marshfield Clinic, Mayo Clinic, Northwestern University, Geisinger Health System, Icahn School of Medicine at Mount Sinai (Mount Sinai), and Group Health Cooperative. Predictors include age, gender, the top principal components (PCs) obtained from the genotypes, as well as single nucleotide polymorphisms (SNPs) that are strongly associated with extreme obesity selected based on p-values from a genome-wide association study (GWAS) and linkage disequilibrium. The screening steps of SNPs are described in the Supplementary Material.

We define populations by race and site, and treat AA at Vanderbilt University as the target population, where a target-only fitting strategy might have limited performance with a sample size of *n* = 646. We include source populations with larger sample sizes than the target population. The sample sizes of the qualified populations are listed in Table 1. Since eMERGE adopts a centralized data sharing model where all individual-level data can be pooled together, we evaluate the centralized transfer learning approach, which is the same as COMMUTE with *M* = 6. In addition, we also evaluate the setting where we assume that individual-level data cannot be shared across sites. In such a case, we have *M* = 1 as data from the Vanderbilt White source population are stored together with the target population.

**Table 1:** Sample size across data centers.

|  |  | Target | Source |  |  |  |  |  |
| --- | --- | --- | --- | --- | --- | --- | --- | --- |
|  |  |  | 1 | 2 | 3 | 4 | 5 | 6 |
|  |  | Vanderbilt University | Vanderbilt University | Marshfield Clinic | Mayo Clinic | Northwestern University | Geisinger Health System | Mount Sinai |
| White | Case | / | 231 | 219 | 142 | 125 | 697 | / |
|  | Total | / | 1523 | 1497 | 1321 | 1305 | 1023 | / |
| African American | Case | 197 | / | / | / | / | / | 235 |
|  | Total | 646 | / | / | / | / | / | 1142 |

The target data are randomly split into training, validation and testing data, with an approximate ratio of 8:2:3. In addition to the target testing dataset, we use the AA samples from Northwestern University (sample size 167) as an external testing dataset which are not included in the training to evaluate the portability of the fitted models. We apply Algorithm 1 with the loss function 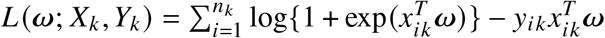 and 𝒫(·) = 𝒬(·) = ∥ · ∥_1_. Over 20 iterations by randomly selecting the training, validation and testing data, we present the result of averaged AUC on the internal and the external testing data, respectively. We compare COMMUTE (with *M* = 1 and *M* = 6) to the target-only, source, and direct aggregation approach introduced in the simulation study.

## Results

### Results of simulation studies

Figures 2 and 3 present AUC over 200 simulation replications over four scenarios, under two different sample size configurations, respectively. In Panels A-C of the two figures, the three source populations have the same *h*_*k*_ (level of heterogeneity). As expected, when the level of heterogeneity increases, the performance of the source estimators on the target data decreases. With low heterogeneity (Panel A), the performance of the source estimators increases with the sample size. With a moderate to high level of heterogeneity, the performance of source estimators may decrease as the sample size grows. With low to moderate heterogeneity (Panels A and B), the source estimators have higher AUC than the target-only estimator, while in the high heterogeneity setting (Panels C), the source estimators have lower AUC than the target-only estimator.

**Figure 2:**
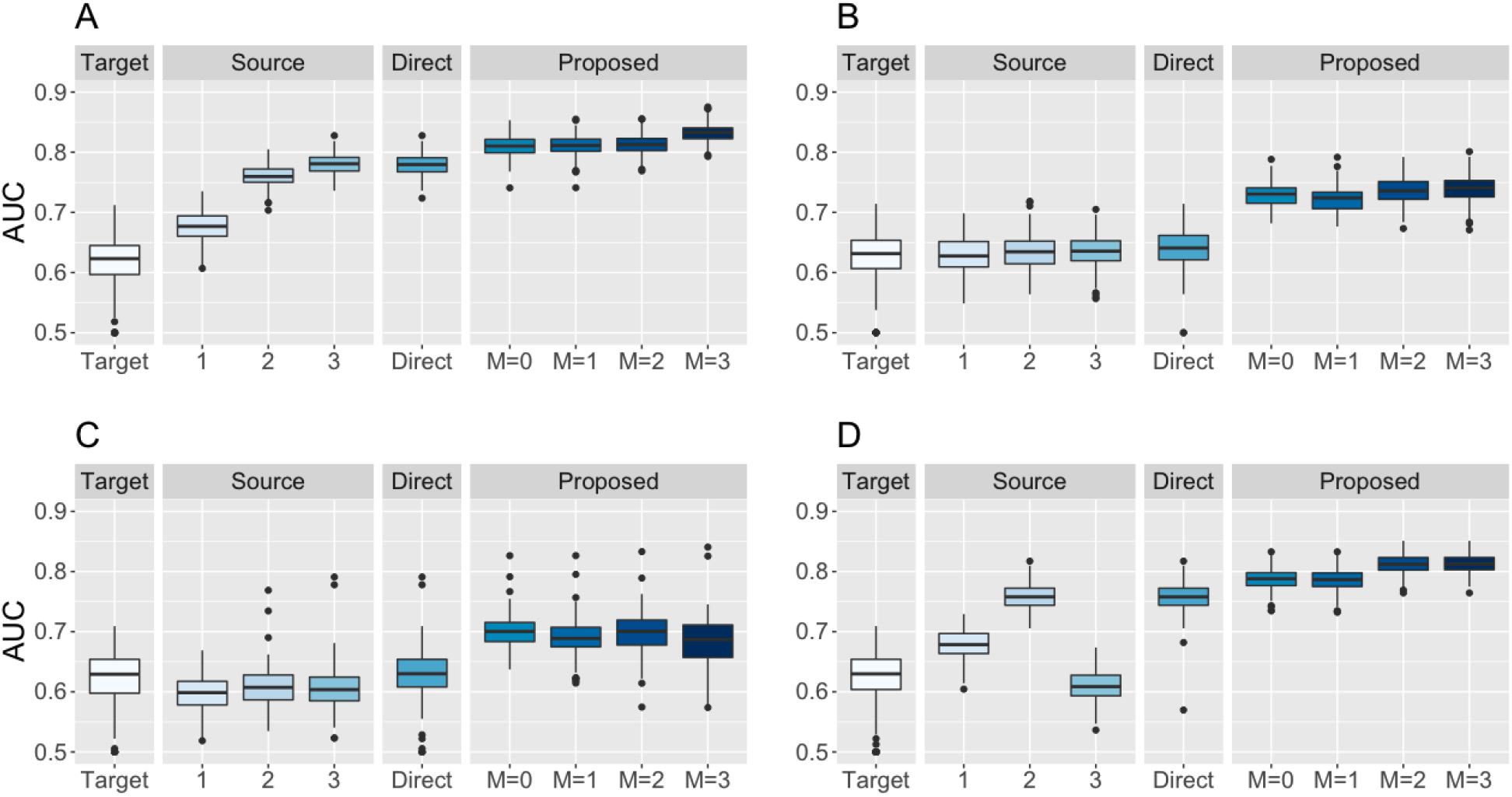
Area under the operating characteristic curve (AUC) over 200 simulation replicates with source sample sizes 1000, 1500, and 2000, respectively. The internal data always has *n* = 500 samples with dimension *p* = 2000. We vary the level of heterogeneity in each source populations with (A) all three source sites have low-level heterogeneity, *h*_1_ = *h*_2_ = *h*_3_ = 20; (B) all three source sites have high level heterogeneity, *h*_1_ = *h*_2_ = *h*_3_ = 120; (C) two source source have low-level heterogeneity, *h*_1_ = *h*_2_ = 20, and one has high level heterogeneity, *h*_3_ = 120; and (D) two source source have low-level heterogeneity, *h*_1_ = *h*_2_ = 20, and one has high level heterogeneity, *h*_3_ = 120. Target: targe-only estimator from equation (1); Source: source estimators from equation (2); Direct: weighted average of target-only and source estimates; Proposed: COMMUTE(federated version, *M* = 0; hybrid version when the first source datasets can be shared with the target, *M* = 1; hybrid version when the first two source datasets can be shared with the target, *M* = 2; centralized version, *M* = 3).

**Figure 3:**
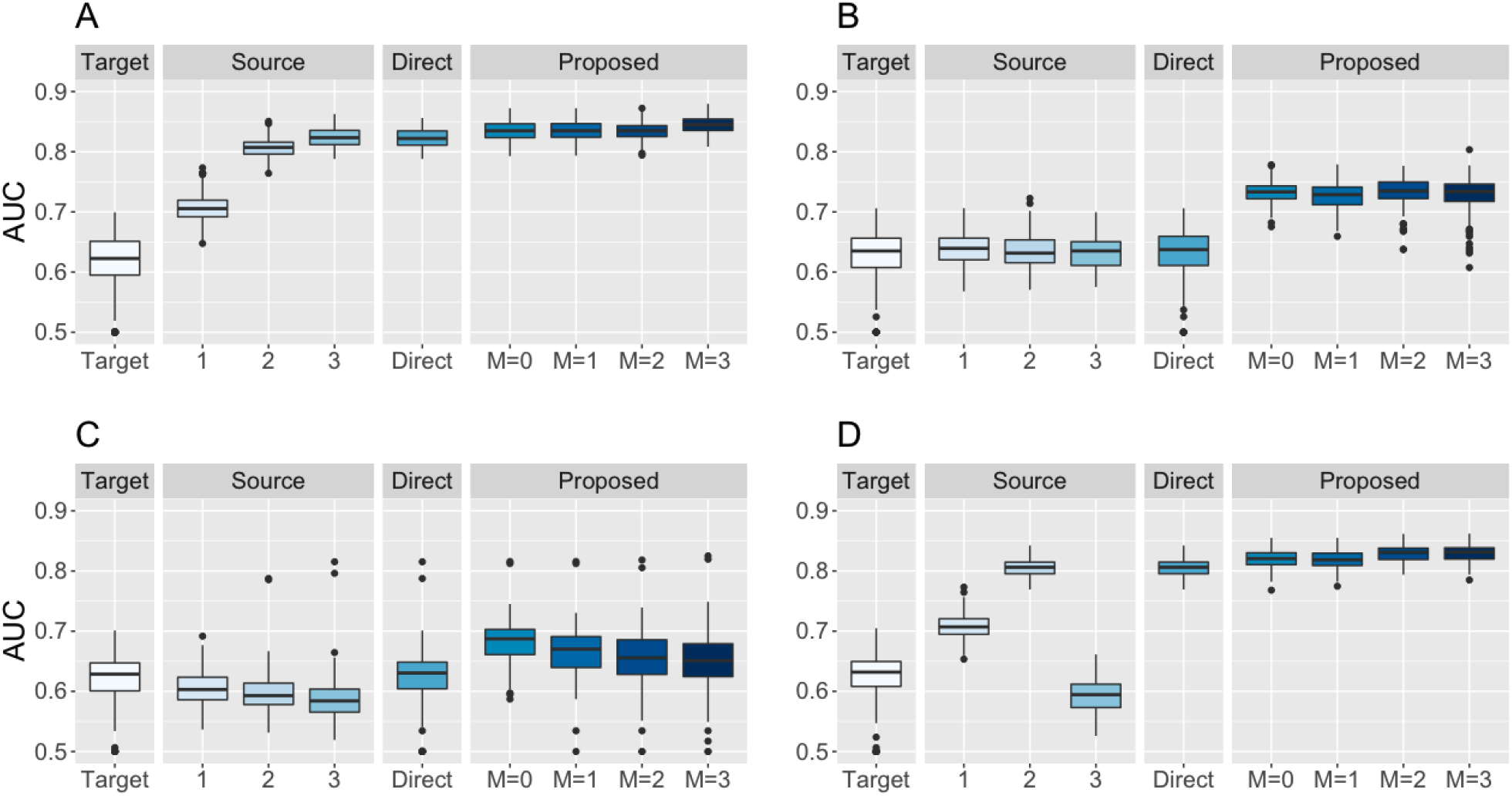
Area under the operating characteristic curve (AUC) over 200 simulation replicates with source sample sizes 2000, 3000, and 4000, respectively. The internal data always has *n* = 500 samples with dimension *p* = 2000. We vary the level of heterogeneity in each source populations with (A) all three source sites have low-level heterogeneity, *h*_1_ = *h*_2_ = *h*_3_ = 20; (B) all three source sites have high level heterogeneity, *h*_1_ = *h*_2_ = *h*_3_ = 120; (C) two source source have low-level heterogeneity, *h*_1_ = *h*_2_ = 20, and one has high level heterogeneity, *h*_3_ = 120; and (D) two source source have low-level heterogeneity, *h*_1_ = *h*_2_ = 20, and one has high level heterogeneity, *h*_3_ = 120. Target: targe-only estimator from equation (1); Source: source estimators from equation (2); Direct: weighted average of target-only and source estimates; Proposed: COMMUTE(federated version, *M* = 0; hybrid version when the first source datasets can be shared with the target, *M* = 1; hybrid version when the first two source datasets can be shared with the target, *M* = 2; centralized version, *M* = 3).

Across all settings, COMMUTE has higher performance than the target-only and source estimators, as well as the direct aggregation approach. The performance of COMMUTE is relatively stable with different *M*. For example, in Panel A of Figure 2, COMMUTE with *M* = 3 has the highest average AUC around 0.83, while COMMUTE with *M* = 0 has AUC of around 0.81. In Panel C, there is a slightly decreasing trend from *M* = 0 to *M* = 3, indicating that having individual-level data does not lead to improved performance than just sharing parameter estimates when the level of heterogeneity is high. Panels C and D show that COMMUTE is robust to the case where some source populations are highly different from the target, and can incorporate information from the source populations that are truly helpful. Comparing Figure 2 to Figure 3, we see that the performance of COMMUTE increases when sample sizes from source populations with low or moderate levels of heterogeneity increase.

### Predicting the risk of extreme obesity using eMERGE data

After screening, 2047 SNPs are included in the model in addition to age, gender and top PCs. For the internal testing at Vanderbilt, as shown in Figure 4, the target-only estimator has an average AUC of around 0.66. The six source models have AUC ranging from 0.50 to 0.63, where the Geisinger model has the worst performance (with AUC of 0.50 indicating no prediction ability on the target data), and the Marshfield model has the best performance on the target population. All of the source estimators have lower performance compared to the target-only estimator, indicating a substantial amount of heterogeneity across populations. The direct aggregation approach which learns a weighted average of the target-only and source estimates shows nearly identical performance compared to the target-only model. The hybrid version of COMMUTE with M=1 has an AUC of around 0.78, comparable to the centralized version of COMMUTE with M=6 (AUC 0.80). Compared to the target-only estimator, COMMUTE has around 12% improvement in AUC. When testing on the external data at Northwestern, the performance of each method is relatively the same as being tested at Vanderbilt. This application has shown that COMMUTE has improved prediction accuracy than the compared methods, and it can efficiently incorporate information from the source when individual-level data can not be shared across sites.

**Figure 4:**
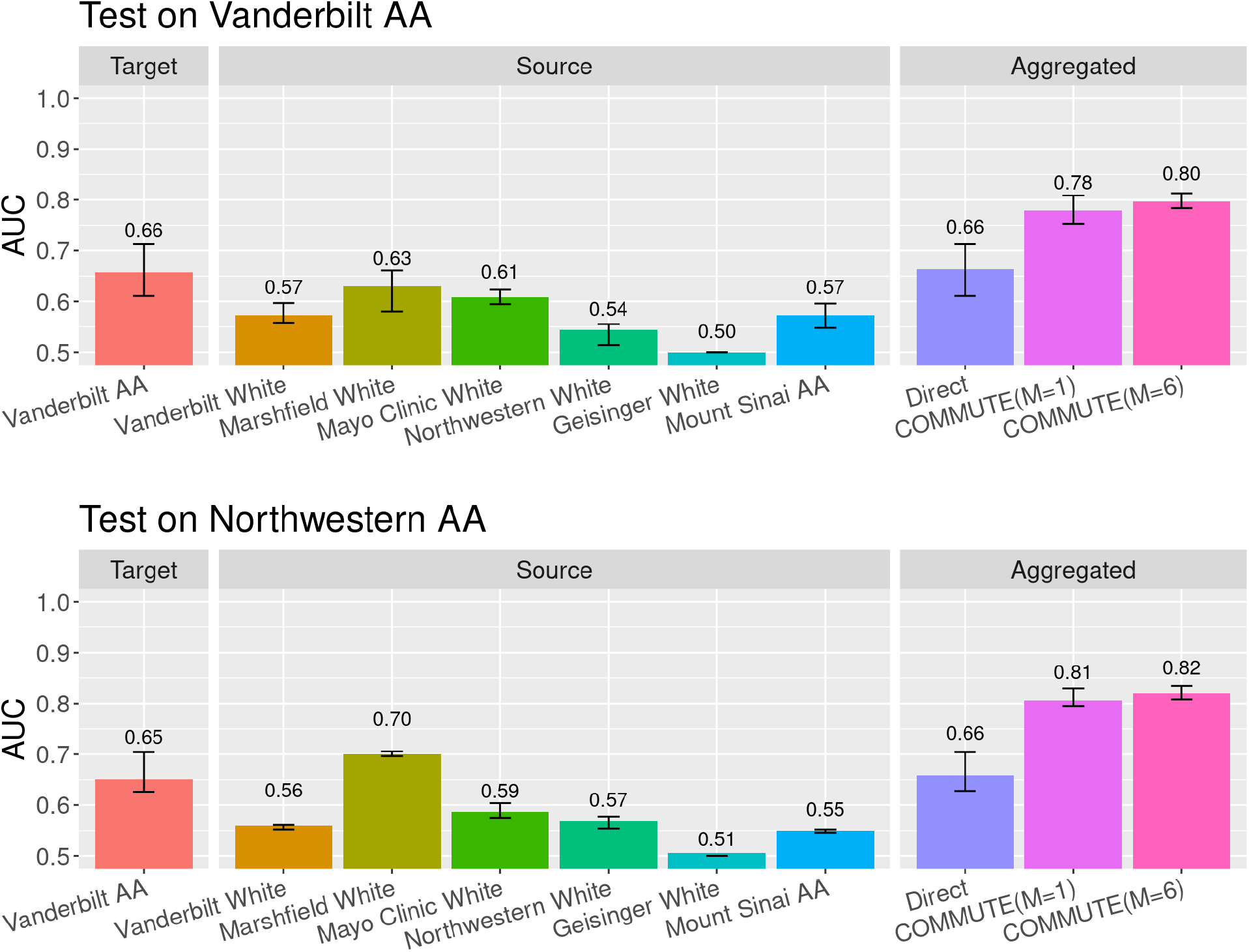
Area under the operating characteristic curve (AUC) of the compared methods for predicting extreme obesity using data from eMERGE. Target: model trained using data from the target population, Vanderbilt African American (AA) only; Source: model trained using data from each source population; Direct: weighted average of the target-only and the source models; COMMUTE: proposed method. Each colored bar shows the average AUC over 10 random replications, the black error bars on top of each colored bar show the 1^st^–3^rd^ quantile of AUC.

## Discussion

In this paper, we propose a transfer learning method, COMMUTE, that can be applied to multisite studies with centralized, federated, or hybrid data sharing models to improve risk prediction in target populations. COMMUTE leverages the similarity across populations to calibrate the source models, generating heterogeneity-adjusted synthetic data corresponding to source data that cannot be directly shared. COMMUTE incorporates available source information to the target data to jointly estimate the model parameters. Our COMMUTE algorithm can safeguard against source populations that are drastically different from the target, and it is communication efficient as each site only needs to share information once with the target site. Through numerical experiments and application to real-world multi-site data, we have illustrated the validity and feasibility of COMMUTE for multi-site risk prediction.

Our method protects data privacy by exchanging only parameter estimates between sites. This level of privacy protection is common in a collaborative research environment, where the risk of membership inference or tracing attacks is low. For example, the Phenotype Knowledgebase website, PheKB, is a collaborative environment for building and validating electronic algorithms to identify characteristics of patients within health data. Researchers are encouraged to share their models trained locally at PheKB to facilitate cross-site collaboration for algorithm development, validation, and sharing for reuse^27^. Moreover, compared to federated algorithms that require iterative sharing of information, such as the gradients of objective functions, COMMUTE has less risk of being attacked by programs that can learn the distribution of individual-level data from the shared gradients^23^. In practice, when necessary, our method can be combined with other privacy protection techniques, such as differential privacy^14,15^ and data encryption^2,62^, to further improve privacy protection. It is worth noticing that some of these approaches may compromise prediction accuracy. Researchers need to balance the level of privacy protection with both the prediction performance and the communication cost in the design of the algorithm.

One limitation of COMMUTE is that it requires all sites to observe the same predictors. In cases where useful predictors for some sites are missing in other sites, the missing features could be first imputed from models trained at other sites^20,36,61^. However, data heterogeneity across sites and populations must be carefully accounted for when applying imputation methods.

COMMUTE is an effective tool to improve prediction performance in a target population. The improvement is more significant when the target population has a smaller sample size than the source populations. This indicates that COMMUTE can help reduce the gaps in the performance of prediction models across populations, which is caused by the limited representation of certain minority and disadvantaged groups. As the current biobank data heavily over-represents European ancestry populations^13,34^, it is a promising and meaningful future direction for us to study the utility of COMMUTE for improving the fairness of risk assessment tools in precision medicine.

## Conclusion

COMMUTE can incorporate models trained from multiple healthcare institutions to improve the performance of risk prediction in a target population. COMMUTE is easy to implement and communication efficient, requiring each site to share only the fitted model parameters. It is robust to the level of heterogeneity across populations. COMMUTE is well-suited to be applied to multisite studies to train robust risk prediction models and address the lack of representation of certain populations.

## Data Availability

The eMERGENetwork data used in this work is publicly available through dbGaP phs000888.v1.p1
(https://ega-archive.org/studies/phs000888).

## Data Availability Statement

TheeMERGENetwork data used in thiswork is publicly available through dbGaP phs000888.v1.p1 (https://ega-archive.org/studies/phs000888).

